# Single time-point multi-dimensional biomarker models can predict acute organ injury trajectory

**DOI:** 10.1101/2025.10.06.25337429

**Authors:** Chris Humphries, Alastair M Kilpatrick, Kathleen M Scullion, Rhona Aird, Lorraine Bruce, Maria Elena Candela, Tak Yung Man, Stuart J Forbes, James W Dear

## Abstract

**Background:** Forecasting acute organ injury trajectory remains a critical clinical challenge. Current approaches rely on serial measurements, delaying decision-making. Using paracetamol-induced liver injury (APAP DILI) as a model, we developed and validated a machine learning framework integrating mechanistically distinct biomarkers to predict injury trajectory from a single timepoint.

**Methods:** We conducted a biomarker discovery and evaluation study using serum from three UK cohorts: MAPP2 APAP DILI biobank discovery cohort (n=160), independent MAIL trial testing cohort (n=34), and healthy controls (n=13). We measured 63 biomarkers and evaluated 321,682 combinations using kernel naïve Bayes classification to predict subsequent ALT trajectory (rising versus falling).

**Findings:** Sensitivity analysis found that model performance and stability were optimized by integrating multiple, weakly-correlated (Spearman ρ<0.5) biomarkers. A seven-biomarker model (CCL5, HMGB1, INR, MCSFR, potassium, sodium, white cell count) predicted injury trajectory with AUC 0.825 (95% CI: 0.685–0.965).

**Interpretation:** This study establishes a generalizable framework demonstrating that robust prognostic models integrate multiple non-collinear biomarkers reflecting distinct biological pathways. This mechanistic triangulation approach provides a systematic route for the translation high-dimensional datasets into clinically tractable tests, with potential applications in sepsis, acute kidney injury, and other acute conditions requiring early patient stratification.

**Funding:** Chief Scientist Office, Scotland (PMAS/21/07); UK Medical Research Council (MR/T044802/1).

## Introduction

Drug-induced liver injury (DILI) remains a major clinical challenge and is the leading cause of acute liver failure in high-income countries, with acetaminophen (APAP) overdose accounting for the majority of cases.^1^ Despite widespread use of N-acetylcysteine (NAC) as the standard antidote, NAC is only effective when administered early and cannot reverse hepatocellular damage once injury has occurred. To date, changes to NAC regimens have primarily impacted tolerability rather than outcomes.^2,3,4^ As a result, efforts to improve outcomes have shifted toward therapies that act beyond the initial insult, targeting inflammation, immune modulation, and regeneration.^5,6^

This has led to the investigation of novel therapeutic strategies that target downstream injury pathways. These range from small molecules that inhibit specific enzymatic pathways, such as fomepizole, to complex interventions like alternatively-activated macrophages (AAMs), a cell therapy currently under evaluation in a phase 1 clinical trial.^7,8 9^

A critical challenge in evaluating these diverse approaches is the reliance on single, imperfect biomarkers. Clinically, the magnitude of liver injury is assessed by measuring blood levels of alanine aminotransferase (ALT). ALT is an enzyme abundant within liver cells (hepatocytes); when these cells are damaged and die, ALT is released into the bloodstream. Its circulating concentration therefore serves as a proxy for the extent of recent hepatocellular necrosis. However, while ALT is a proxy for cell death, it provides little information about specific biological processes (such as inflammation, immune modulation, and regeneration) that these novel therapies aim to influence. The in vivo behaviour of biomarkers reflecting these targeted mechanisms remains poorly characterised, limiting our ability to stratify patients, monitor therapeutic response, or identify those most likely to benefit.

To date, most biomarker research in APAP drug-induced liver injury (APAP DILI) has focused on individual markers and uses threshold-based or cross-sectional comparisons.^10^ While these approaches have been useful, they overlook the multi-dimensional nature of liver injury biology and the potential power of combining complementary, non-redundant biological signals. There is a pressing need for a more nuanced understanding of biomarker behaviour across the injury time-course and between phenotypes, as well as improved methods to interpret complex, multi-marker datasets. This challenge is not unique to DILI; it is a common hurdle in many dynamic disease states, such as acute kidney injury or sepsis, where single biomarkers often fail to capture the complete pathophysiological picture.^11,12^

While the potentially fatal consequence of APAP DILI is acute liver failure (ALF), clinical decisions around antidote selection, escalation to novel therapeutics, or conservative management are ideally made much earlier, during the broader evolving phase of APAP DILI.^13^ Improving biomarker resolution in this phase has the potential to guide critical decisions about who to treat, when to escalate, and when to withhold therapy as novel interventions emerge. The ultimate goal of treatment is to prevent progression to ALF, and the greatest opportunity to intervene is by identifying patients on a worsening trajectory early.^14^ This would allow novel treatments to be targeted to those with the greatest potential to benefit, rather than relying on crude ALT thresholds or waiting for clinical deterioration.

In this study, we conducted a multiplex biomarker analysis in patients with APAP overdose to address these challenges. We aimed to map a broad landscape of biomarker behaviour across injury phenotypes and time; apply dimensionality reduction and multivariate analysis techniques to uncover patterns not captured by ALT alone; and identify novel individual and combination biomarkers that forecast the trajectory of liver injury.

By shifting the focus from a single proxy measure of established organ injury like ALT to the multi-dimensional biology of liver injury, we aimed to establish a methodological framework for developing predictive models that may be generalizable to other forms of acute organ injury.

## Methods

### Design and setting

This was a biomarker discovery and evaluation study conducted using human serum samples from three distinct UK-based cohorts. The overall process is summarised in Fig. 5. Initial model discovery utilised samples selected from the multi-centre MAPP2 biobank. Model performance was evaluated on an independent testing cohort of pre-intervention samples collected from participants in the ongoing Macrophages for Acute Liver Injury (MAIL) trial; ISRCTN12637839. A third cohort of healthy donor samples, obtained from the University of Edinburgh, was used to establish biomarker reference ranges. All APAP patients received routine clinical care.

### Study cohorts and sample processing

Discovery human APAP DILI serum samples were selected from APAP DILI survivors in the MAPP2 biobank based on peak alanine aminotransferase (ALT) values (collected July 2018-May 2023). Three patient phenotypes were defined based on the peak ALT reached by that patient: no ALT rise (peak ALT <50 U/L), sub-hepatotoxic DILI (peak ALT 50-999 U/L), and hepatotoxic DILI (peak ALT ≥1000 U/L). All samples from a given patient were assigned to that patient’s phenotype, regardless of the ALT at the specific timepoint. Thirteen healthy donor plasma samples were obtained to establish reference ranges by calculating median-adjusted deviation (MAD) Z-scores. This method calculates a Z-score for each discovery biomarker by measuring its deviation from the median of the healthy donor samples. Unlike standard Z-scores this approach minimises the influence of extreme outliers, providing a more reliable measure of how far a patient’s biomarker level deviates from the healthy reference range. Samples for the independent testing cohort were provided from participants in the MAIL trial (pre-intervention only; collected Oct 2023-Apr 2025). All samples were obtained after the development of APAP DILI hepatotoxicity.

Patients with less severe APAP DILI are typically discharged from hospital at earlier timepoints following overdose. As our samples were obtained from a biobank reflecting routine clinical care, this resulted in baseline differences in the time period studied in each cohort. This was addressed analytically by grouping samples by overall patient phenotype for some analyses, while specifically examining temporal biomarker profiles in others.

As this was an exploratory biomarker discovery study, a formal a priori sample size calculation was not performed, a protocol was not prepared, and study-specific patient and public involvement was not undertaken. The size of the testing cohort was determined by the number of eligible participants with appropriate samples available from the MAIL trial. The study was designed to include the maximum number of available samples to explore a wide range of biomarker signals and to provide an independent dataset for initial model testing.

### Biomarker quantification

Potential biomarkers were selected through a combination of literature review, local consensus, multiplex platform availability, and available serum volumes. A full list of all 63 biomarkers and their assays is provided separately (Supplementary Information S6).

To explore the broadest range of potential novel biomarkers in each sample, model discovery samples were analysed in singlicate, with assay standards in duplicate, in an approach consistent with European Bioanalysis Forum recommendations.^15^ Testing samples were analysed in duplicate. All assays were undertaken by technicians blinded to sample phenotypes through the use of pseudonymisation codes. Sample aliquoting and de-pseudonymisation was undertaken by a non-blinded study team member.

### Statistical and bioinformatic analysis

#### Data pre-processing and management

Where volume permitted, all analyses were undertaken on *n*=160 samples, with the exception of Luminex cytokines (*n*=159), HMGB1 ELISA (*n*=159), and miR-122 RT PCR (*n*=159). Where values were reported as unreadable due to analyser failure, they were removed from analysis (*n*=1 for CCL5, MCSFR, and PDGF-BB). Assays for CX3CL1 and CXCL14 were removed from analysis because the majority of measurements fell on the lower plateau of the standard curve, indicating poor quantitative resolution.

Biomarkers with significant proportions of censored data (outside of limits of quantification) could not have missing values imputed, as assumptions regarding underlying data distribution could not be made. While values beyond the limits of quantification could be treated as categorical data, this approach is not compatible with the downstream quantitative and multivariate analyses central to this study. Therefore, assays demonstrating data censoring in ≥50% of samples (CCL8, IL-1a, IL-1b, IL-3, IL-4, IL-21, IL-23) were considered to have insufficient quantitative resolution for this exploratory study and were excluded from all analyses. For remaining biomarkers (*n*=44), censored data were substituted with the lowest or highest value within the limits of detection as appropriate, with the addition of a small constant (e.g. -0.1 or +0.1) to maintain rank values.

For all subsequent dimensionality reduction and predictive modelling analyses, missing values were imputed using the kNN function from the VIM package with k=5. For final model development focused on translatable biomarkers, imputation was re-performed after restricting the dataset to the 15 biomarkers available in both the discovery and testing cohorts. Imputation was required for 0.8% of the data points in the final discovery matrix. No imputation was needed for the testing matrix.

#### Modelling and statistics

All statistical and bioinformatic analyses were performed using R (v4.4.2). Group comparisons used a Kruskal-Wallis, followed by post-hoc Dunn’s tests with Bonferroni correction for significant (p<0.05) biomarkers. Temporal relationships between each biomarker and ALT decline were assessed in patients with serial samples post-ALT peak, categorising the biomarker’s decline as ‘Earlier’, ‘Later’, or the ‘Same’ relative to ALT.

For dimensionality reduction, data were scaled and centred. A Spearman correlation matrix was computed and visualised using corrplot (v0.95). Principal Component Analysis (PCA) was performed via eigendecomposition of the Spearman matrix. Uniform Manifold Approximation and Projection (UMAP) was also performed using the uwot package (v0.2.3, with n_neighbours=15, min_dist=0.2; all other parameters default).

Supervised models were built using a kernel naïve Bayes classifier from the e1071 R package (v1.7-14) to classify subsequent ALT trajectory (rising vs. falling, on the next blood test obtained as part of routine clinical care). To select uncorrelated features, we first performed a sensitivity analysis to identify an optimal Spearman correlation threshold. For the final models, an exhaustive search identified all biomarker combinations (size 1-8) that met the correlation threshold. Model performance was estimated using 5-fold cross-validation, with Area Under the ROC Curve (AUC) as the primary metric, using the pROC package (v1.19.0.1). The top 30 performing models for each size were trained on the full discovery dataset and then tested on the independent cohort. AUC in the testing cohort and 95% confidence intervals were calculated using 2000 bootstrap replicates.

### Ethics and funding

Collection and analysis of human samples from the MAPP2 biobank was approved by the London South East Research Ethics Committee (18/LO/0894). Collection and analysis of healthy donor samples obtained through the Centre for Inflammation Research Blood Donor Register, University of Edinburgh, was approved by the Edinburgh Medicine and Veterinary Medicine Research Ethics Committee (21-EMREC-041). Testing samples were provided from pre-intervention serum obtained in the ongoing MAIL Trial (Macrophages Therapy for Acute Liver Injury), a clinical trial approved by North East (York) Research Ethics Committee (reference 23/NE/0019), NHS Lothian Research and Development department and the MHRA. This study was funded by the Centre for Precision Cell Therapy for the Liver (PRaCTicaL), a Chief Scientist Office funded research centre (PMAS/21/07), and via the Medical Research Council funded MAIL Trial (MR/T044802/1). Funders had no role in writing the manuscript or the decision to submit for publication. No payment was received for writing this article. Authors were not precluded from accessing data in the study and accept responsibility to submit for publication. For the purpose of open access, the author has applied a Creative Commons Attribution (CC BY) licence to any Author Accepted Manuscript version arising from this submission.

## Results

### Baseline characteristics

**Table 1.** Baseline characteristics of patients in the discovery cohort. ALT = alanine aminotransferase (U/L), APAP = acetaminophen, DILI = Drug-induced liver injury, IQR = interquartile range.

| Patient phenotype | Healthy control | No DILI<br>(Peak ALT < 50) | Sub-hepatotoxic DILI<br>(Peak ALT 50-999) | Hepatotoxic DILI<br>(Peak ALT ≥ 1000) |
| --- | --- | --- | --- | --- |
| Number of patients | 13 | 15 | 13 | 20 |
| Total number of samples | 13 | 45 | 28 | 74 |
| Sample time range since final APAP ingestion (hrs) | N/A | 0.5-50.1 | 2.0-82.0 | 12.7-154.6 |
| Peak ALT during admission (U/L) (median, IQR) | N/A | 20<br>(17-23) | 122<br>(112-195) | 3736<br>(2473-5645) |
| Median ALT of samples (U/L) (median (IQR)) | 32<br>(30-40) | 16<br>(13-20) | 146<br>(105-337) | 2255<br>(1288-3653) |

Three DILI phenotypes were defined, based on the peak ALT concentration observed during patient admission: no DILI (in which ALT remained less than the upper limit of normal), sub-hepatotoxic DILI (in which a rise in ALT was evident, but remained sub-hepatotoxic), and hepatotoxic DILI (peak ALT ≥ 1000). This upper threshold was chosen as it is a widely accepted indicator of significant, acute hepatocellular injury.^16^

### Characterisation of novel biomarker profiles in APAP DILI

We studied a panel of biomarkers that measure hepatocellular injury, cell signalling, organ function and broader metabolic state. Exploratory analysis of the biomarker landscape across our cohort used a heatmap to visualise patterns stratifying by injury severity and time-course (Fig. 1). Multiple biomarkers showed dynamic changes, highlighting distinct biological responses across APAP DILI phenotypes.

**Fig. 1.**
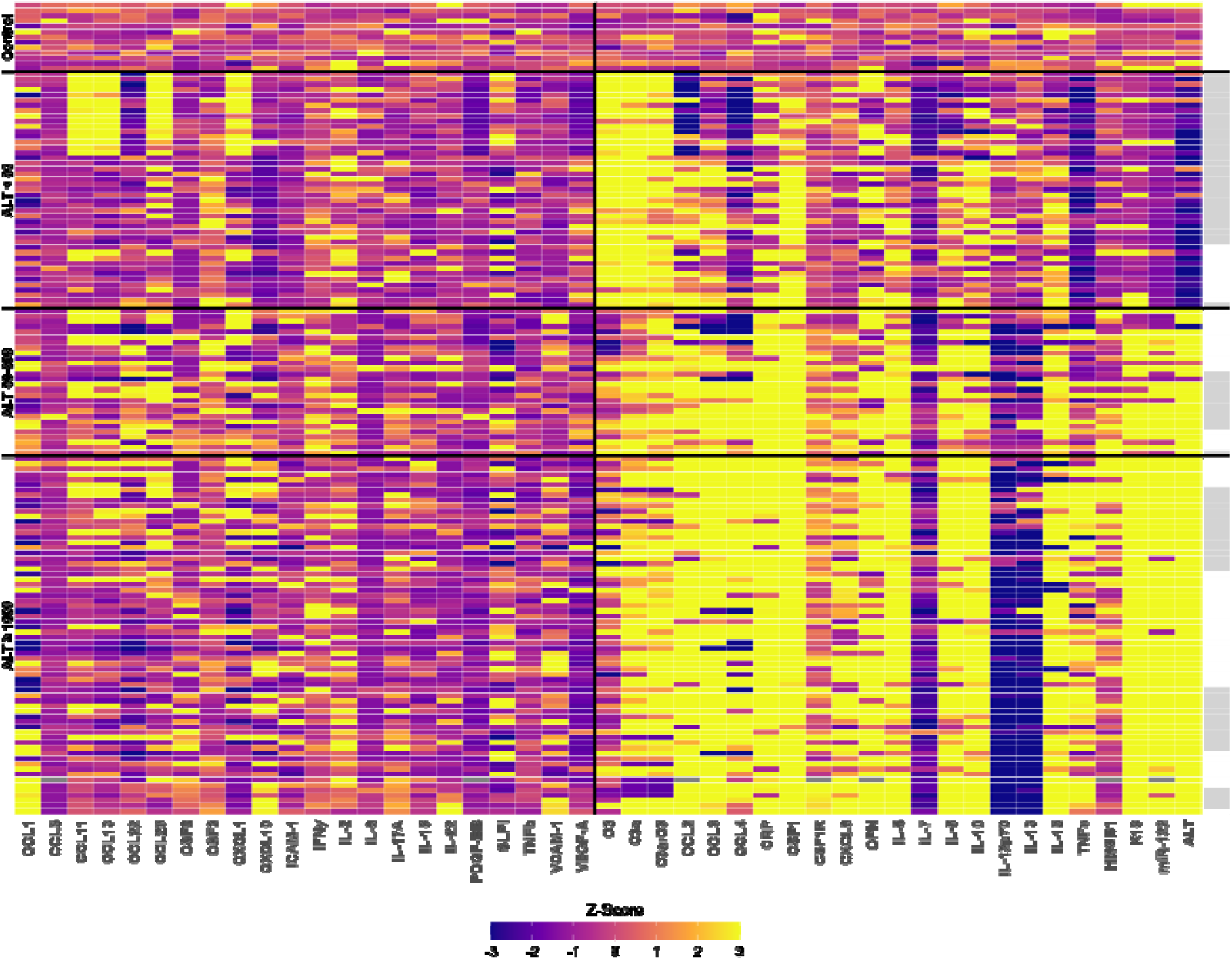
Novel biomarker expression following paracetamol ingestion, stratified by injury phenotype. Heatmap of novel biomarker values. Values are expressed in n=154 samples as median adjusted deviation Z-scores derived from control values. Biomarker phenotype (far left) describes the peak ALT value reached by patients within that phenotype group. Within each phenotype, samples are ordered according to the time since last reported APAP ingestion. Alternating white and grey bars on the right of the figure indicate 24hr periods from last APAP ingestion within each group. To highlight different patterns of response, biomarkers are grouped based on the magnitude of change: those with largely stable or inconsistent change in concentrations versus those with distinct evidence of change in APAP DILI. n=6 ALT ≥ 1000 phenotype samples are not shown as there was no reported time of last ingestion.

Biomarkers associated with hepatocyte injury (such as keratin-18 (K18) and microRNA-122 (miR-122)) largely mirrored the behaviour of ALT, rising in parallel with increasing liver damage. Some biomarkers exhibited more complex patterns. For example, Complement C3 and its activated fragment C3a were elevated in patients with APAP overdose without DILI but were notably depleted early in DILI.

Several cytokines and chemokines showed patterns that correlated with injury severity. Notably, CCL2, CCL3, CCL4, IL-6, IL-8, and IL-10 appear to be elevated in proportion to DILI severity, while IL-12p70 and IL-13 decreased with more severe injury. Colony stimulating factor 1 (CSF1) demonstrated particularly compelling changes across phenotypes, potentially reflecting macrophage recruitment and activation.

### Biomarkers distinguish clinical phenotypes of injury severity

Having established that multiple biomarkers are altered following APAP overdose, we next examined whether they could differentiate between our defined clinical phenotypes (no ALT rise, sub-hepatotoxic, and hepatotoxic). For these comparisons, all samples from a given patient were grouped into that patient’s final peak ALT phenotype, regardless of the ALT value at the individual sampling timepoint.

This analytical approach was chosen to identify biomarkers that reflect the patient’s overall biological trajectory and predisposition to severe injury, rather than those that simply correlate with the contemporaneous ALT concentration. For example, this method could uncover markers that are elevated early in patients destined for severe injury, even when their ALT is still low.

Eleven biomarkers did not reach statistical significance using Kruskal-Wallis testing. The remaining biomarkers proceeded to pairwise post hoc comparisons, with Benjamini-Hochberg-corrected p-values presented in Fig. 2-A.

**Fig. 2:**
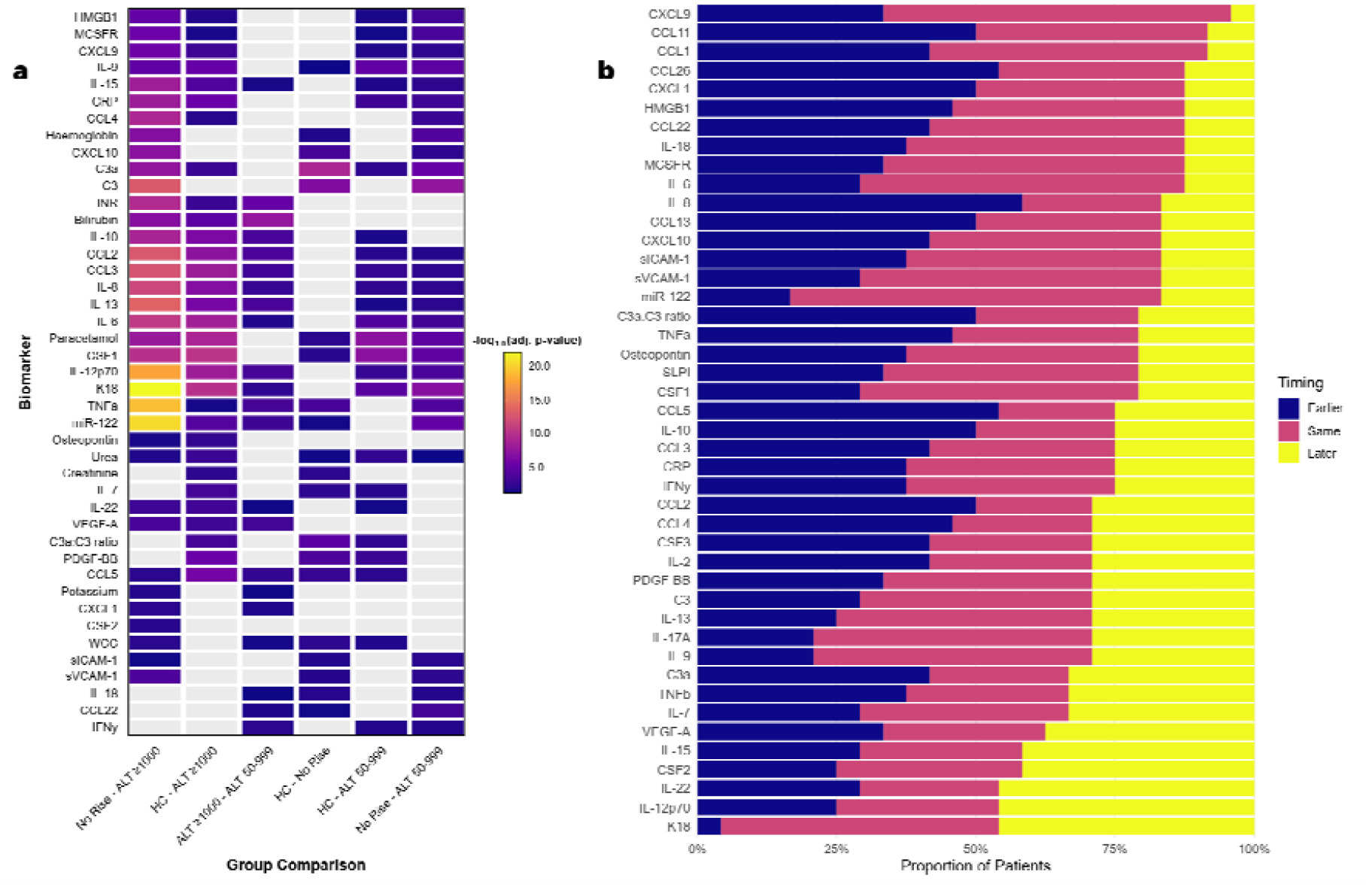
Phenotypic and kinetic differences in biomarkers. A) Biomarkers with a statistically significant difference between phenotype groups obtained by Kruskal-Wallis. (p<0.05). Results of post-hoc comparison tests with adjusted p<0.05 shown in colour. Results are ordered by hierarchical clustering. B) Stacked bar chart showing the relative timing of biomarker decline compared to ALT among patients with an identifiable ALT peak (n = 24). Bars represent the proportion of samples in which each biomarker fell earlier than ALT (“wins”), later than ALT (“losses”), or in the same sampling interval (“equal”). Biomarkers are ordered by ascending number of losses. As samples were collected during routine clinical care, time intervals are coarse; simultaneous decline is pragmatically classified as “equal” despite the potential for small timing differences.

A subset of biomarkers robustly distinguished the hepatotoxic phenotype from less severe groups. These included markers of inflammation (e.g. IL-6, IL-10), cell death (K18, miR-122), and liver function (INR, bilirubin). Many of these were able to discriminate between all three APAP DILI phenotypes (sub-hepatotoxic DILI, hepatotoxic DILI, and no DILI), while showing no significant difference between healthy controls and non-DILI overdose patients, suggesting potential utility in the stratification of DILI severity.

Several biomarkers—such as C3a, the C3a:C3 ratio, CCL5, and PDGF-B—showed significant differences between all APAP-exposed phenotypes and healthy controls, regardless of peak ALT. These may be indicative of sub-clinical liver injury, early biological response prior to detectable hepatocellular damage, or associated with paracetamol exposure itself.

Other biomarkers (CRP, CXCL9, HMGB1, MCSFR) consistently differentiated between liver injury (ALT rise) and uninjured patients (healthy controls and non-DILI), but did not distinguish between sub-hepatotoxic and hepatotoxic injury. These markers may therefore reflect a more binary transition from no injury to injury, rather than tracking injury severity.

Together, these findings demonstrated that many biomarkers change with injury phenotype, and that these relationships may reflect distinct biological processes underpinning the onset, progression, and severity of APAP-induced liver injury.

### Biomarker time-course profiles differ from ALT in acute liver injury

Recognising that comparisons by phenotype included samples from varying times post-overdose, we next focused on whether individual biomarkers might decline earlier than ALT during recovery, potentially allowing earlier identification of improving liver injury.

We analysed patients from DILI cohorts (peak ALT ≥50 IU/L) who had at least one prior sample available for comparison, yielding 79 samples from 24 individuals. For each biomarker, we assessed whether its concentration fell before, after, or at the same time as ALT (Fig. 2-B). The outcome was the frequency with which each biomarker declined earlier than ALT), declined later, or showed equal performance.

No individual biomarker consistently won or lost; a weighted ‘win ratio’ ((‘fell before’-’fell later’)/all) suggested that IL-8, CCL26 and CCL11 fell earlier than ALT most often.

These results demonstrate that many biomarkers are likely to exhibit time-course patterns that differ from ALT. Although no individual biomarker consistently fell earlier than ALT across all patients, the variability in kinetics suggested that refining the cohort in whom a single biomarker is measured, or combining biomarkers within predictive models have the potential to offer earlier insights into liver injury trajectory.

### Relationships between biomarkers inform redundancy and complementarity

We next examined the relationships between biomarkers to assess potential redundancy or complementarity. Strongly correlated biomarkers often reflect a shared biological origin, such as the concurrent release of ALT, K18, and miR-122 during hepatocyte necrosis, and consequently may provide limited added value when combined in a predictive model.^11^ In contrast, biomarkers with weak or no correlation may describe distinct biological pathways.

To explore this, we computed pairwise correlations between biomarkers in patients with peak ALT ≥50 (Figure 3). Subsequent clustering indicated groups of biomarkers that showed significant and moderate to strong correlations with each other, particularly among those reflecting hepatocyte injury (e.g., ALT, K18, miR-122). However, many other biomarkers demonstrated weak pairwise correlations, including no significant correlation with ALT itself.

**Fig. 3:**
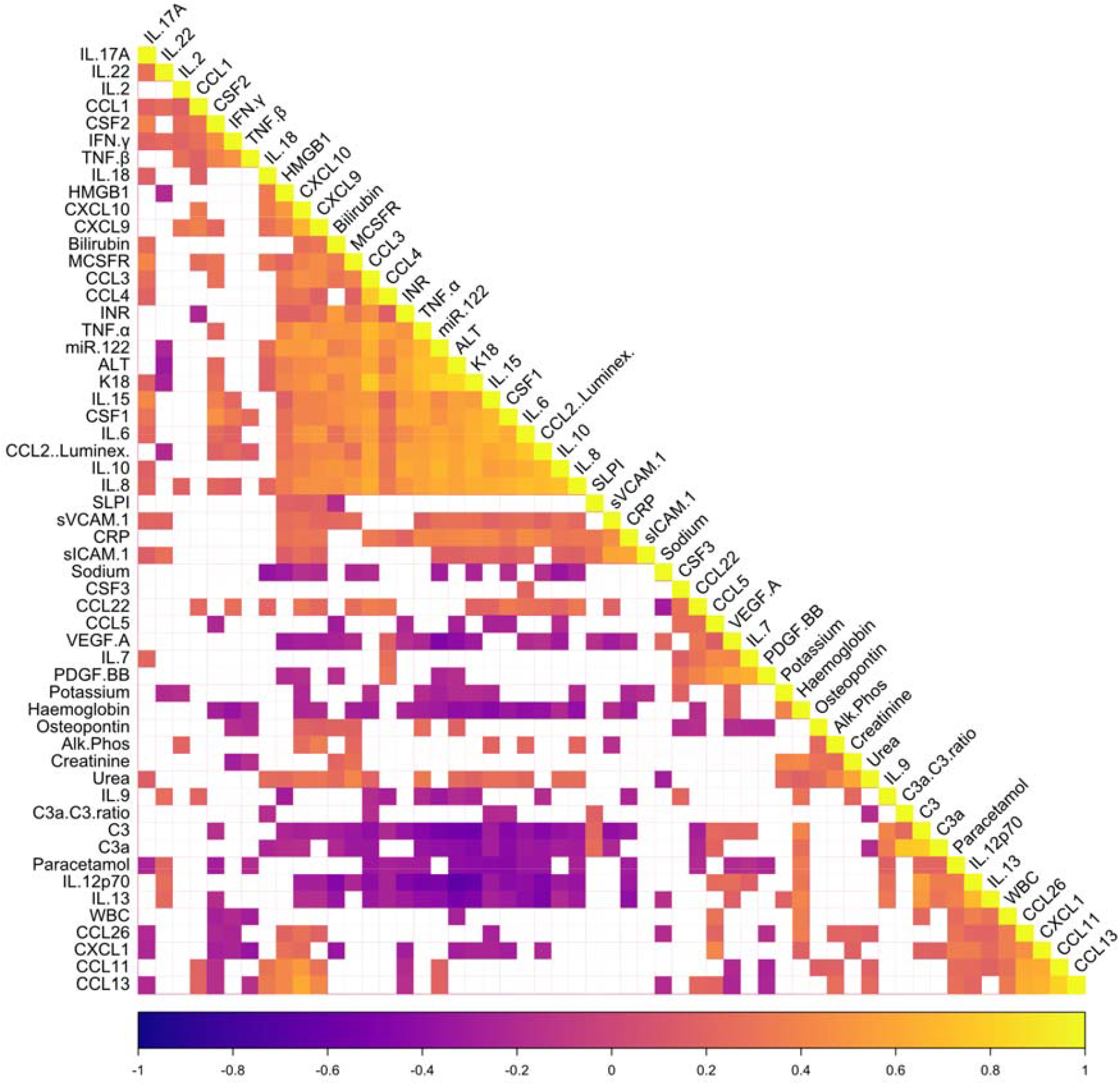
Correlogram of all analysed biomarkers. Biomarkers are clustered by correlation; only significant correlations (p<0.05) are shown, and are colour coded by the strength of correlation as assessed by Spearman’s rank correlation.

This lack of strong correlation, even among biomarkers that were individually associated with injury phenotype or severity, suggests that magnitude alone does not explain biomarker behaviour. Instead, differences in temporal profiles and pathophysiological origin may allow certain biomarkers to provide complementary information to ALT; particularly when predicting future trajectory rather than capturing current injury state.

These findings support the use of multi-dimensional biomarker models, where uncorrelated or partially correlated biomarkers can be leveraged to improve prediction of organ injury trajectory. While routine clinical biomarkers are essential for identifying and monitoring liver injury, the added value of novel biomarkers in contextualising why and how this injury is developing is essential for improved predictive power.

### Dimensionality reduction captures phenotypic clustering, but not injury trajectory

Having established that individual biomarkers vary in magnitude and time-course, and have the potential to provide complementary data, we examined whether a dimensional reduction approach considering all biomarkers (for example, via Principal Component Analysis or UMAP) could distinguish clinical phenotypes and predict injury trajectory (Supplemental Information S3).

Dimensionality reduction successfully separated patients based on injury severity (Supplementary Fig. S3-3AB). However, these approaches failed to identify injury trajectory, even within the hepatotoxic group (Supplementary Fig. S3-3CD). Crucially, this finding demonstrates that while biomarker concentrations reflect the state of injury, the signals predicting its future course are more subtle and complex than can be resolved by unsupervised clustering alone.

This prompted a more direct, supervised approach. Direct comparison between ALT-rising and ALT-falling samples confirmed that multiple biomarkers differed significantly between the two trajectories (Supplemental Table S3-4), motivating the development of a predictive model.

### Multi-dimensional biomarker models predict injury trajectory in APAP DILI

#### Concept exploration

Given that unsupervised methods failed to resolve injury trajectory (Supplementary Fig. S2-3), we developed a supervised machine learning approach. Using samples obtained following the development of hepatotoxicity (ALT > 1000 IU/L), we trained a kernel naïve Bayes classification model to predict whether the subsequent ALT measurement would be rising or falling.

ALT alone was a poor predictor of its own trajectory, with an area under the receiver operating characteristic curve (AUC) of 0.625 (95% CI: 0.441–0.810). In contrast, several individual novel biomarkers performed better, with IL-10 and CCL5 achieving the highest AUCs (both 0.783; Supplementary Table S4-1). To assess whether combining mechanistically-distinct signals improved prediction, we tested pairwise combinations of biomarkers with low inter-correlation (ρ<0.7, chosen as values above this threshold are known to distort modelling).^17^ This approach improved performance substantially, with 68 biomarker pairs outperforming the best single biomarker (Supplementary Table S4-1), supporting the concept that developing multi-dimensional models could improve predictive power. An example of the incremental gains to AUC is provided as supplemental Fig. S4-2.

#### Model discovery

To derive the optimal biomarker combination, we balanced predictive power against the risk of overfitting. We performed an exhaustive search for models containing one to four biomarkers and used a targeted beam search for larger models (n=378,658 potential models, see Supplemental Information S4). This revealed progressively smaller gains in performance beyond four biomarkers (Supplementary Fig. 4-3A). Sensitivity analysis identified that restricting model components to biomarkers with a maximum correlation of ρ<0.5 optimized model performance in the testing cohort (Supplementary Information S5), balancing biomarker collinearity against restricting the number of potential models.

Having established this new correlation threshold, we re-derived our models, with the beam search expanded to n=1000 (*n*=321,682 models). This suggested that reducing biomarker correlation thresholds results in improvements at model discovery as more biomarkers are added (Fig. 4-A).

**Fig. 4:**
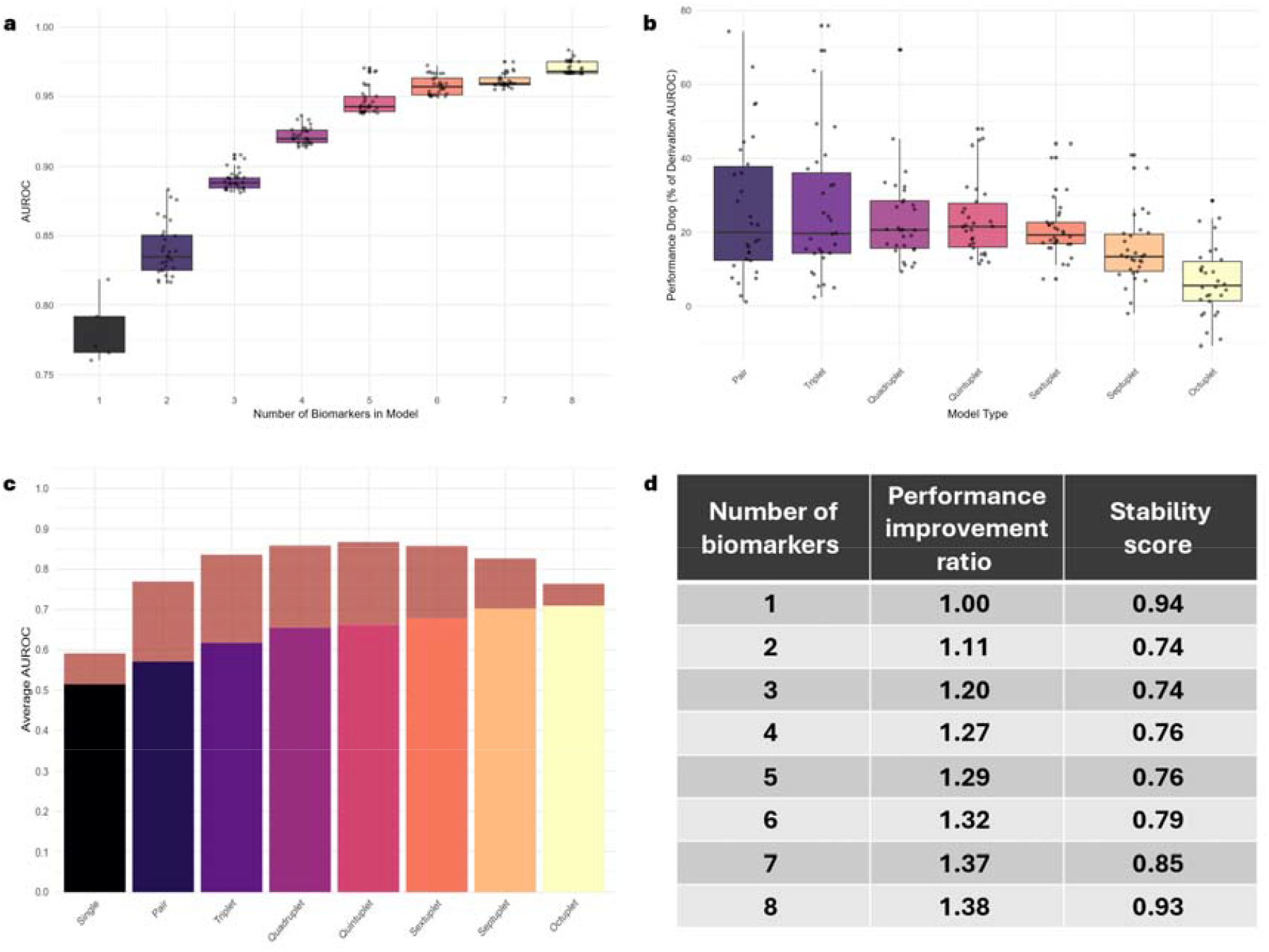
Discovery and testing of multi-dimensional models. A) Boxplot of the top 30 discovery AUCs for each model size. B) Box-plot showing the impact of model size on translation from discovery to testing datasets for the top 30 models at each combination model size, indicating increased robustness as additional biomarkers are added. C) Stacked bar chart showing the mean performance of the top 30 biomarker models from 1-8 biomarkers at testing and the mean performance drop from discovery. D) Statistics for the mean performance of the top 30 biomarker models showing performance improvement ratios [(testing AUROC for n biomarkers)/(testing AUROC for 1 biomarker)] and stability scores [1 - (mean % drop from discovery to testing)] demonstrating that increasing the number of mechanistically distinct biomarkers leads to increased performance, and that this improvement is increasingly stable at testing with larger models.

**Fig. 5:**
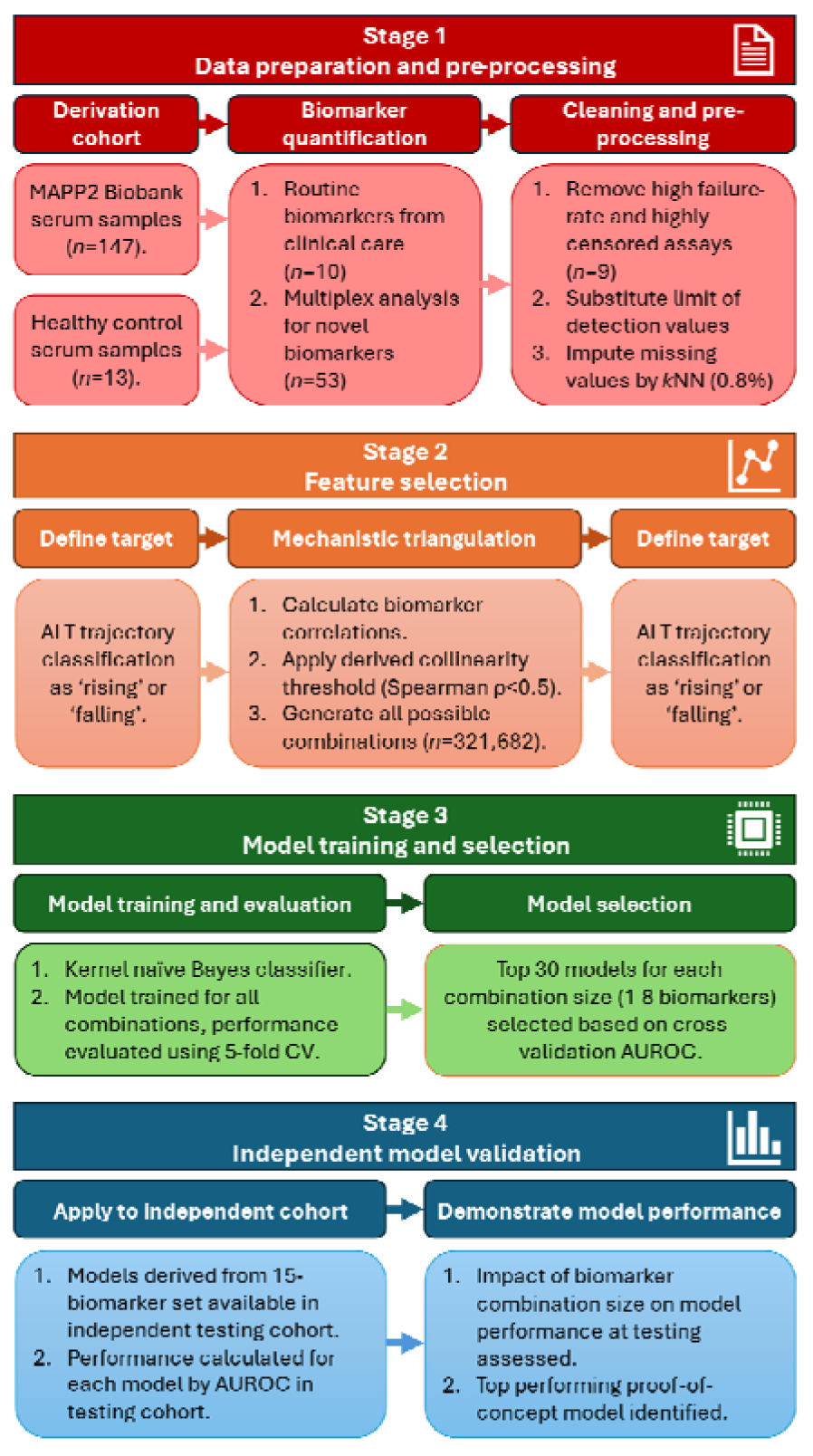
Study flow diagram. Abbreviations: ALT = Alanine Aminotransferase, AUROC = Area Under the Receiver-Operating Characteristic curve, CV = Cross-Validation, *k*NN = *k*-Nearest Neighbour imputation.

#### Model testing in an independent cohort

We tested our models in an independent testing cohort of *n*=34 samples from patients with APAP-induced hepatotoxicity (*n*=10 rising, n=24 falling). Testing was restricted to models built from *n*=15 biomarkers quantified on identical platforms across both cohorts (Supplementary Information S4.3). It was not possible to produce sufficient models for analysis at biomarker combinations larger than 8, within the correlation threshold restrictions.

In this testing cohort, we found that increasing numbers of mechanistically-distinct biomarkers consistently improved model stability, with smaller falls in performance observed in the testing set (Fig. 4-B). Increasing numbers of biomarkers within models consistently improved the mean performance of the top 30 models (Fig. 4-C). The single top-performing model from the restricted biomarker set available in the testing cohort was a seven-biomarker combination - CCL5, HMGB1, INR, MCSFR, Potassium, Sodium and WCC - which achieved an AUC of 0.825 (95% CI: 0.685–0.965).

Mechanistically, this seven-biomarker model integrates biomarkers of inflammation (CCL5), liver necrosis (HMGB1), impaired liver synthetic function (INR), macrophage activation (MCSFR), the systemic immune response (WCC), and broad metabolic dysregulation (potassium and sodium), likely providing a more holistic assessment of the patient’s pathophysiological state.

## Discussion

Predicting the trajectory of acute organ injury is a fundamental challenge across medicine.^18^ Current options are limited to serial assessment of biomarkers, which delays decision-making; prognostic tools like the King’s College Criteria for acute liver failure, which are only applied once organ failure is established and thus limit the ability to apply preventive therapies; or clinical scoring systems which, as exemplified by the challenges in sepsis, often have poor predictive performance for individual patient trajectories.^19,20^ In this study, we use acetaminophen-induced liver injury (APAP DILI) as a well-characterised model to address this challenge. We demonstrate that by moving beyond a single, dominant biomarker and using multidimensional models, we can predict whether injury is dynamically worsening or resolving.

Our most significant finding is the development and independent testing of a model that predicts the future trajectory of liver injury from a single timepoint. The current standard, a single ALT measurement, was a poor predictor of its own future direction (AUC 0.625). In contrast, our top-performing 7-biomarker model achieved an AUC of 0.825, representing a substantial improvement in predictive power.

This result was underpinned by key methodological insights. We found that multi-dimensional biomarkers models can effectively integrate multiple weakly predictive signals to forecast injury trajectory, and that the path to generating robust and stable predictive models lies in the careful selection of biomarkers to minimize collinearity (ρ<0.5). This principle of ‘mechanistic triangulation’, integrating weaker but distinct biological signals, was more effective than relying on a few dominant, but correlated, markers, providing a clear rationale for why biomarker models which do not explicitly account for collinearity require rigorous external testing due to the risk of overfitting.^16^ The approach followed provides a foundational template for developing multi-analyte predictive models from any complex dataset, and aligns with the broader shift in medicine from single-biomarker ‘magic bullets’ to a systems-based understanding of disease pathophysiology.^21^

We propose that the success of our 7-biomarker model is attributable to its ability to capture the complex pathophysiology of APAP DILI. Rather than relying on a single aspect of injury, the panel provides a holistic snapshot of the patient’s state by integrating signals of inflammation (CCL5), hepatocyte necrosis (HMGB1), impaired liver function (INR), macrophage activation (MCSFR), the systemic immune response (WCC), and broad metabolic disturbance (potassium and sodium).^22,23,24^ This integrated assessment explains why the model can predict a dynamic outcome like trajectory, where unsupervised methods failed. The model succeeds not because any single marker is dominant, but because it learns the optimal weighting of these diverse signals to determine whether the patient’s biological state is tilting towards further injury or recovery.

The development of accurate, single-timepoint predictive tools is a pressing clinical priority in acute care.^25^ Such tools would enable a paradigm shift from reactive monitoring to proactive decision-making, optimizing the allocation of limited resources like intensive care unit (ICU) beds and guiding the timely deployment of targeted interventions. This is particularly crucial for the development of novel therapies. Clinical trials for acute illnesses are frequently confounded by patient heterogeneity, which can obscure a true treatment effect.^26^ A validated predictive model, such as the one proposed here, could serve as a patient enrichment tool, enabling smaller, more statistically powerful, and efficient trials.^27^ Although our 7-marker model requires validation in larger, prospective cohorts, it serves as a robust proof-of-concept for this predictive methodology.

The broad applicability of our findings stems from the development of a generalizable methodological framework. The challenge of forecasting patient trajectory with single biomarkers is common to many acute conditions, including acute kidney injury, pancreatitis, and sepsis. This is particularly relevant for the growing number of academic and industrial biomarker discovery consortia that generate vast, high-dimensional datasets. Such groups often face a significant bottleneck in translating these discoveries into clinically tractable tests. Our work provides a systematic roadmap to navigate this challenge: 1) profiling of a broad, mechanistically diverse biomarker panel; 2) feature selection based on a pre-defined collinearity threshold to prioritize complementary signals; 3) systematic modelling of biomarker combinations; and 4) external validation. This principle of ‘mechanistic triangulation’ therefore represents a strategy to progress from large-scale discovery data to robust, mechanistically interpretable, and clinically relevant predictive tools.

A key strength of this study is the use of well-phenotyped clinical samples across the full spectrum of APAP DILI, analysed using a broad, blinded-analysis biomarker panel. A further significant strength is the testing of our predictive modelling approach in a separate patient cohort, which increases confidence in the robustness of our findings.

However, the study has limitations. The initial discovery phase is exploratory, and the analysis of multiple biomarkers introduces a risk of Type I error. While our primary modelling approach was evaluated in an independent cohort, this testing cohort itself was small; therefore, further validation in larger, more diverse patient populations is required. The reliance on samples from routine clinical care also resulted in coarse sampling intervals, and our cohorts were restricted to survivors, meaning signatures may differ in ALF. Finally, plasma biomarker levels remain an indirect measure of the hepatic microenvironment.

In conclusion, our study suggests that multi-dimensional biomarker signatures can forecast the trajectory of acute organ injury. Using APAP DILI as a model, we developed and tested a simple probabilistic classifier that can forecast the trajectory of liver injury from a single timepoint by combining uncorrelated biomarkers representing distinct biological processes.

More broadly, our work provides a tested, generalizable framework for developing predictive models in other complex disease states. The approach of combining mechanistically-distinct, low-collinearity biomarkers to forecast trajectory could be readily applied to other conditions featuring acute organ injury. By providing a window into the in vivo pathophysiology, this methodology has the potential to accelerate the development of targeted treatments and improve our understanding of acute organ injury across multiple clinical contexts.

## Supporting information

checklist

## Acknowledgements

We thank K. Wilson at the University of Edinburgh SURF assay facility for support with assays, and all investigators on the MAIL Trial for their participation in clinical trial delivery which supported this study. The authors acknowledge financial support from the Medical Research Council (reference MR/T044802/1) and the Chief Scientist’s Office Scotland via the Center for Precision Cell Therapy for the Liver (PMAS/21/07). UKRI MRC, CSO Scotland.

## Data availability

The authors declare that all data supporting the findings of this study are available within the paper and its supplementary information files. Requests for further data and analytical code may be addressed to the corresponding author.

## Contributions

CH conceived the study concept and experimental design. CH, AMK, KMS, RA, LB, MEC, TYM performed experiments. CH, AMK analysed the data. CH, AMK, JWD provided intellectual input and supervision. CH drafted the manuscript. CH, AMK, SJF, JWD reviewed and edited the manuscript.

## References

1. Vento, Sandro et al. Acute liver failure in low-income and middle-income countries. The Lancet Gastroenterology & Hepatology, Volume 8, Issue 11, 1035–1045

2. Humphries C, Clarke E, Eddleston M, Gillings M, Irvine S, Keating L, Miell A, Milne L, Muir L, O’Brien R, Oatey K, Raman R, Thanacoody R, Tuck S, Weir CJ, Wood DM, Dear JW; HiSNAP Trial Investigators. HiSNAP trial-a multicentre, randomised, open-label, blinded end point, safety and efficacy trial of conventional (300 mg/kg) versus higher doses of acetylcysteine (450 mg/kg and 600 mg/kg) in patients with paracetamol overdose in the UK: study protocol. BMJ Open. 2025 Mar 22;15(3):e097432. doi: 10.1136/bmjopen-2024-097432. PMID: 40122550; PMCID: PMC11977480.

3. Humphries C, Pettie J, Agboola B, Caparrotta TM, Hunter RW, Morrison E, Sandilands EA, Webb DJ, Eddleston M, Dear J. Scottish and Newcastle Antiemetic Protocol (SNAP) 12-hour acetylcysteine regimen for paracetamol overdose reduces anaphylactoid reactions without compromising hepatic protection in all age groups: a secondary analysis. Emerg Med J. 2025 Aug 24:emermed-2024-214533. doi: 10.1136/emermed-2024-214533.

4. Humphries C, Roberts G, Taheem A, Abdel Kader H, Kidd R, Smith J. SNAPTIMED study: does the Scottish and Newcastle Antiemetic Protocol achieve timely intervention and management from the emergency department to discharge for paracetamol poisoning? Emerg Med J. 2023 Mar;40(3):221–223. doi: 10.1136/emermed-2021-212180. Epub 2022 Aug 18. PMID: 35981856.

5. Woolbright BL, Jaeschke H. Role of the inflammasome in acetaminophen-induced liver injury and acute liver failure. J Hepatol. 2017 Apr;66(4):836–848. doi: 10.1016/j.jhep.2016.11.017. Epub 2016 Nov 29.

6. Humphries C, Addison ML, Dear JW, Forbes SJ. The emerging role of alternatively activated macrophages to treat acute liver injury. Arch Toxicol. 2025 Jan;99(1):103–114. doi: 10.1007/s00204-024-03892-2. Epub 2024 Nov 6.

7. Humphries C, Addison M, Aithal G, et al. Macrophage Therapy for Acute Liver Injury (MAIL): a study protocol for a phase 1 randomised, open-label, dose-escalation study to evaluate safety, tolerability and activity of allogeneic alternatively activated macrophages in patients with paracetamol-induced acute liver injury in the UK. BMJ Open. 2024 Dec 9;14(12):e089417. doi: 10.1136/bmjopen-2024-089417.

8. Starkey Lewis P, Campana L, Aleksieva N, et al. Alternatively activated macrophages promote resolution of necrosis following acute liver injury. J Hepatol. 2020 Aug;73(2):349–360. doi: 10.1016/j.jhep.2020.02.031. Epub 2020 Mar 11.

9. Candela ME, Addison M, Aird R, et al. Cryopreserved human alternatively activated macrophages promote resolution of acetaminophen-induced liver injury in mouse. NPJ Regen Med. 2025 Jan 22;10(1):5. doi: 10.1038/s41536-025-00393-3.

10. Humphries C, Dear JW. Novel biomarkers for drug-induced liver injury. Clin Toxicol (Phila). 2023 Aug;61(8):567–572. doi: 10.1080/15563650.2023.2259089. Epub 2023 Oct 10. PMID: 37767912.

11. Dear JW, Clarke JI, Francis B, Allen L, Wraight J, Shen J, Dargan PI, Wood D, Cooper J, Thomas SHL, Jorgensen AL, Pirmohamed M, Park BK, Antoine DJ. Risk stratification after paracetamol overdose using mechanistic biomarkers: results from two prospective cohort studies. Lancet Gastroenterol Hepatol. 2018 Feb;3(2):104–113. doi: 10.1016/S2468-1253(17)30266-2. Epub 2017 Nov 14. PMID: 29146439; PMCID: PMC5777094.

12. Woolbright BL, Jaeschke H. Mechanisms of Inflammatory Liver Injury and Drug-Induced Hepatotoxicity. Curr Pharmacol Rep. 2018 Oct;4(5):346–357. doi: 10.1007/s40495-018-0147-0. Epub 2018 Jun 30. PMID: 30560047; PMCID: PMC6294466.

13. Fontana RJ, Liou I, Reuben A, et al. AASLD practice guidance on drug, herbal, and dietary supplement-induced liver injury. Hepatology. 2023 Mar 1;77(3):1036–1065. doi: 10.1002/hep.32689.

14. Cardoso FS, Lee WM, Karvellas CJ; US Acute Liver Failure Study Group. Failure to rescue in acute liver failure: A multicenter cohort study. Liver Transpl. 2025 Aug 1;31(8):982–988. doi: 10.1097/LVT.0000000000000594.

15. Barfield M, Goodman J, Hood J, Timmerman P. European Bioanalysis Forum recommendation on singlicate analysis for ligand binding assays: time for a new mindset. Bioanalysis. 2020 Mar;12(5):273–284. doi: 10.4155/bio-2019-0298. Epub 2020 Jan 24.

16. Yau CE, Chen H, Lim BP, Ng M, Ponampalam R, Lim DYZ, Chin YH, Ho AFW. Performance of the paracetamol-aminotransferase multiplication product in risk stratification after paracetamol (acetaminophen) poisoning: a systematic review and meta-analysis. Clin Toxicol (Phila). 2023 Jan;61(1):1–11. doi: 10.1080/15563650.2022.2152350.

17. Dormann C, Elith J, Bacher S, et al. Collinearity: A review of methods to deal with it and a simulation study evaluating their performance. Ecography. 2013;36(1):27–46. doi: 10.1111/j.1600-0587.2012.07348.x

18. Vincent JL, Moreno R. Clinical review: scoring systems in the critically ill. Crit Care. 2010;14(2):207. doi: 10.1186/cc8204. Epub 2010 Mar 26.

19. O’Grady JG, Alexander GJ, Hayllar KM, Williams R. Early indicators of prognosis in fulminant hepatic failure. Gastroenterology. 1989 Aug;97(2):439–45. doi: 10.1016/0016-5085(89)90081-4.

20. Seymour CW, Liu VX, Iwashyna TJ, et al. Assessment of Clinical Criteria for Sepsis: For the Third International Consensus Definitions for Sepsis and Septic Shock (Sepsis-3). JAMA. 2016 Feb 23;315(8):762–74. doi: 10.1001/jama.2016.0288. Erratum in: JAMA. 2016 May 24-31;315(20):2237. doi: 10.1001/jama.2016.5850.

21. Barabási AL, Gulbahce N, Loscalzo J. Network medicine: a network-based approach to human disease. Nat Rev Genet. 2011 Jan;12(1):56–68. doi: 10.1038/nrg2918.

22. Li M, Sun X, Zhao J, et al. CCL5 deficiency promotes liver repair by improving inflammation resolution and liver regeneration through M2 macrophage polarization. Cell Mol Immunol. 2020 Jul;17(7):753–764. doi: 10.1038/s41423-019-0279-0.

23. Liu J, Jiang M, Jin Q, et al. Modulation of HMGB1 Release in APAP-Induced Liver Injury: A Possible Strategy of Chikusetsusaponin V Targeting NETs Formation. Front Pharmacol. 2021 Jul 21;12:723881. doi: 10.3389/fphar.2021.723881. PMID: 34366873; PMCID: PMC8333615.

24. Church RJ, Kullak-Ublick GA, Aubrecht J, et al. Candidate biomarkers for the diagnosis and prognosis of drug-induced liver injury: An international collaborative effort. Hepatology. 2019 Feb;69(2):760–773. doi: 10.1002/hep.29802. Epub 2018 Jun 27.

25. Maslove DM, Tang B, Shankar-Hari M, et al. Redefining critical illness. Nat Med. 2022 Jun;28(6):1141–1148. doi: 10.1038/s41591-022-01843-x.

26. Sinha P, Calfee CS. Phenotypes in acute respiratory distress syndrome: moving towards precision medicine. Curr Opin Crit Care. 2019 Feb;25(1):12–20. doi: 10.1097/MCC.0000000000000571.

27. U.S. Food and Drug Administration. Enrichment Strategies for Clinical Trials to Support Approval of Human Drugs and Biological Products. 2019. Available from: https://www.fda.gov/regulatory-information/search-fda-guidance-documents/enrichment-strategies-clinical-trials-support-approval-human-drugs-and-biological-products [accessed Sep 2025].

