## supplemental for "Single time-point multi-dimensional biomarker models can predict acute organ injury trajectory"

**Supplemental Information**

#### S1. Quantification of CCL2 across multiplex platforms

The correlation between Mesoscale Discovery and Luminex platforms for the quantification of CCL2 as assessed by Spearman’s ρ was 0.881 (95% CI 0.840 – 0.913, p<0.001), though absolute quantification values were noted to be different.

**Fig. S1-1: Multiplex platform comparison.**

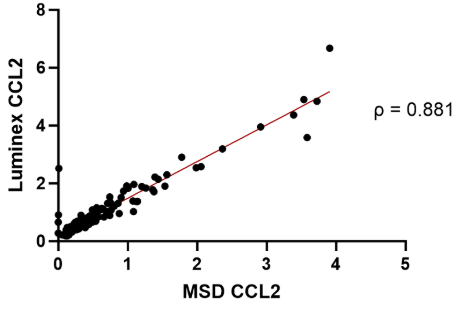

**Correlation of quantification of chemokine ligand 2 (CCL2, ng/mL) quantified by Luminex Discovery Assay and Mesoscale Discovery V-PLEX (Human) Assay.**

#### S2. Prediction of hepatotoxicity

To assess whether individual biomarkers could predict hepatotoxicity before it was evident by ALT, we selected samples obtained within the first 48 hours of reported paracetamol ingestion where ALT was still <1000 IU/L. Samples were classified into two groups: those from patients who subsequently developed hepatotoxicity (ALT ≥1000; n=15) and those who did not (n=67).

Among 46 biomarkers tested, 21 showed statistically significant differences between these groups. These were subjected to ROC curve analysis, and AUCs with 95% confidence intervals were compared to ALT. These results are shown in Table S2-1.

While ALT performed extremely well in this predictive analysis (AUC = 0.924), this is likely inflated by spectrum bias introduced by the selection criteria, limiting inclusion to patients with ALT <1000. In this group, those who ultimately developed hepatotoxicity were often already in the early stages of ALT rise, even if below the diagnostic threshold.

This enriched separation creates an optimistic AUC for ALT that would not translate directly into clinical practice where ALT values and trajectories are more heterogeneous. As a result, and given the limited additional value that could be demonstrated beyond ALT in this context, we did not pursue multi-dimensional biomarker modelling or validation.

However, the relative performance of novel biomarkers within this analysis still supports the prioritisation of candidate biomarkers for future clinical validation in true early diagnostic settings.

| **Biomarker** | **p value** | **AUC** |
| --- | --- | --- |
| **CCL4** | ***<0.001*** | ***0.939*** |
| **ALT** | ***<0.001*** | ***0.924*** |
| **miR-122** | ***<0.001*** | ***0.914*** |
| **CCL3** | ***<0.001*** | ***0.889*** |
| **K18** | ***<0.001*** | ***0.863*** |
| **TNFa** | ***<0.001*** | ***0.859*** |
| **IL-8** | ***<0.001*** | ***0.829*** |
| **IL-10** | ***<0.001*** | ***0.797*** |
| **CCL2 MSD** | ***<0.001*** | ***0.788*** |
| **CCL2 Lum** | ***<0.001*** | ***0.787*** |
| IL-6 | 0.004 | 0.734 |
| HMGB1 | 0.005 | 0.728 |
| **IL-13** | ***0.007*** | ***0.722*** |
| IL-9 | 0.008 | 0.706 |
| CRP | 0.010 | 0.711 |
| CSF1 | 0.012 | 0.706 |
| C3 | 0.013 | 0.704 |
| IL-22 | 0.017 | 0.696 |
| IL-12p70 | 0.017 | 0.696 |
| C3a | 0.024 | 0.686 |
| VEGF-A | 0.049 | 0.664 |

**Table S2-1. Biomarker values with a statistically significant difference in samples from patients who subsequently develop hepatotoxicity, and those who do not. AUCs for which the 95% confidence intervals overlap with ALT (0.868—0.989) are displayed in *bold*.**

**Fig. S2-2: Prediction of hepatotoxicity**

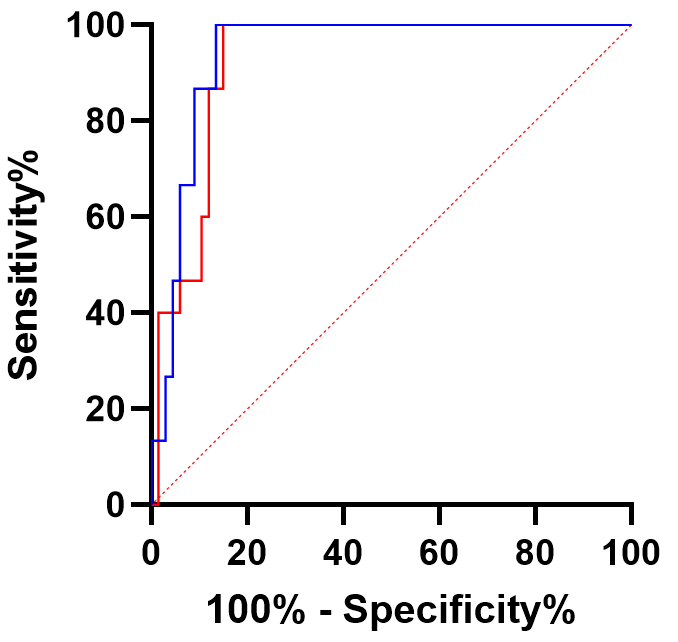

**ROC curve of CCL4 (blue) and ALT (red) obtained from patients without hepatotoxicity in the first 48 hours following last ingestion of paracetamol, for the prediction of subsequent hepatotoxicity (ALT ≥1000).**

#### S3. Dimensionality reduction fails to predict ALT trajectory

###### PCA analysis

We conducted a semi-supervised principal component analysis (PCA) using a Spearman’s correlation matrix to explore the full APAP dataset (n=147) and assess whether samples clustered by clinical phenotype without knowledge of ALT values. After loading and cleaning the dataset, we performed k-nearest neighbour imputation for missing values and standardized the biomarker data. PCA revealed distinct groupings of APAP overdose patients without liver injury, and those with DILI, suggesting that multi-dimensional biomarker profiles can distinguish clinically relevant subgroups (Fig. S3-3A). This supports the potential utility of novel biomarkers for the early stratification of patients by injury severity.

However, PCA failed to separate samples based on ALT trajectory (i.e. whether ALT was rising or falling at the time of sampling), limiting its ability to distinguish worsening vs. recovering liver injury. Projection into UMAP space showed a similar pattern: DILI samples and non-injury samples formed clusters, but there was no meaningful separation between ALT-rising and ALT-falling samples (Fig. S3-3B), indicating the need to find specific predictors for ALT trajectory.

Principal component 1 (PC1) accounted for 21.8% of variance in the data and was strongly correlated with ALT in post hoc analysis (ρ = 0.812, p = 2.2 × 10^⁻16^), suggesting it reflects overall injury burden. However, no biomarker dominated the PC1 loadings (Table S3-1), indicating that many biomarkers have the potential to inform prediction of injury burden. The top 10 biomarkers were all noted to have co-localised with ALT in our correlogram.

| **Biomarker** | **PC1 loading** |
| --- | --- |
| K18 | 0.259011 |
| miR-122 | 0.236312 |
| TNFa | 0.233774 |
| IL-8 | 0.231844 |
| CSF1 | 0.231542 |
| IL-6 | 0.228319 |
| CCL2 | 0.227507 |
| CCL3 | 0.225166 |
| IL-10 | 0.218273 |
| IL-15 | 0.215862 |

**Table S3-1. Top 10 biomarkers associated with PC1, ordered by PCA loading.**

Given the clinical importance of distinguishing patients with worsening vs. improving liver injury, we next asked whether multi-dimensional biomarker data could predict ALT trajectory from a single timepoint sample (i.e., whether the subsequent ALT would rise or fall). We first analysed all APAP samples (n=147) and found that biomarkers appeared to distinguish phenotypes (supplement 1).

Focussing on hepatotoxicity, we repeated our analyses in the peak ALT >1000U/L group. PCA and UMAP again showed no clear separation between ALT-rising and ALT-falling groups (Fig. XXXG-H), indicating that dimensionality reduction alone does not resolve trajectory within this subset. However, direct comparisons using Mann-Whitney testing identified sixteen potentially useful biomarkers that significantly differed between the two groups, suggesting that triangulation of multiple biomarker signals may allow discrimination where low-dimensional projections fail to separate classes. These findings prompted exploration of predictive modelling approaches capable of integrating multiple non-correlated weakly informative features to classify ALT trajectory.

###### Biomarker differences by injury trajectory

Using the APAP dataset (n=147), we compared biomarker levels between ALT-rising and ALT-falling samples using Kruskal-Wallis and post hoc Dunn's tests. Four novel biomarkers (CCL22, IL-7, CCL4, CCL5) were significantly lower in ALT-falling samples (Fig. S2-3 E-H). Notably, CCL22 was significantly higher in ALT climbing than in non-DILI or ALT falling groups, while IL-7 and CCL5 saw the same pattern for ALT falling. CCL4 saw significant differences between all groups. These findings indicated that individual biomarkers have the potential to inform ALT trajectory prediction in individual samples, and that there is the potential to use these biomarkers in combination to increase discriminatory potential.

| **Biomarker** | **Kruskal-Wallis p-value** | **Pairwise p-value (adj.)** |
| --- | --- | --- |
| CCL22 | 7.33E-06 | 0.000579 |
| IL-7 | 0.000128 | 0.001988 |
| WCC | 0.000296 | 0.00567 |
| CCL4 | 2.68E-10 | 0.006875 |
| CCL5 | 0.00231 | 0.008213 |

**Table S3-2. Biomarkers found to be significantly different between ALT climbing and ALT falling phenotypes in all APAP DILI samples, ordered by p-value.**

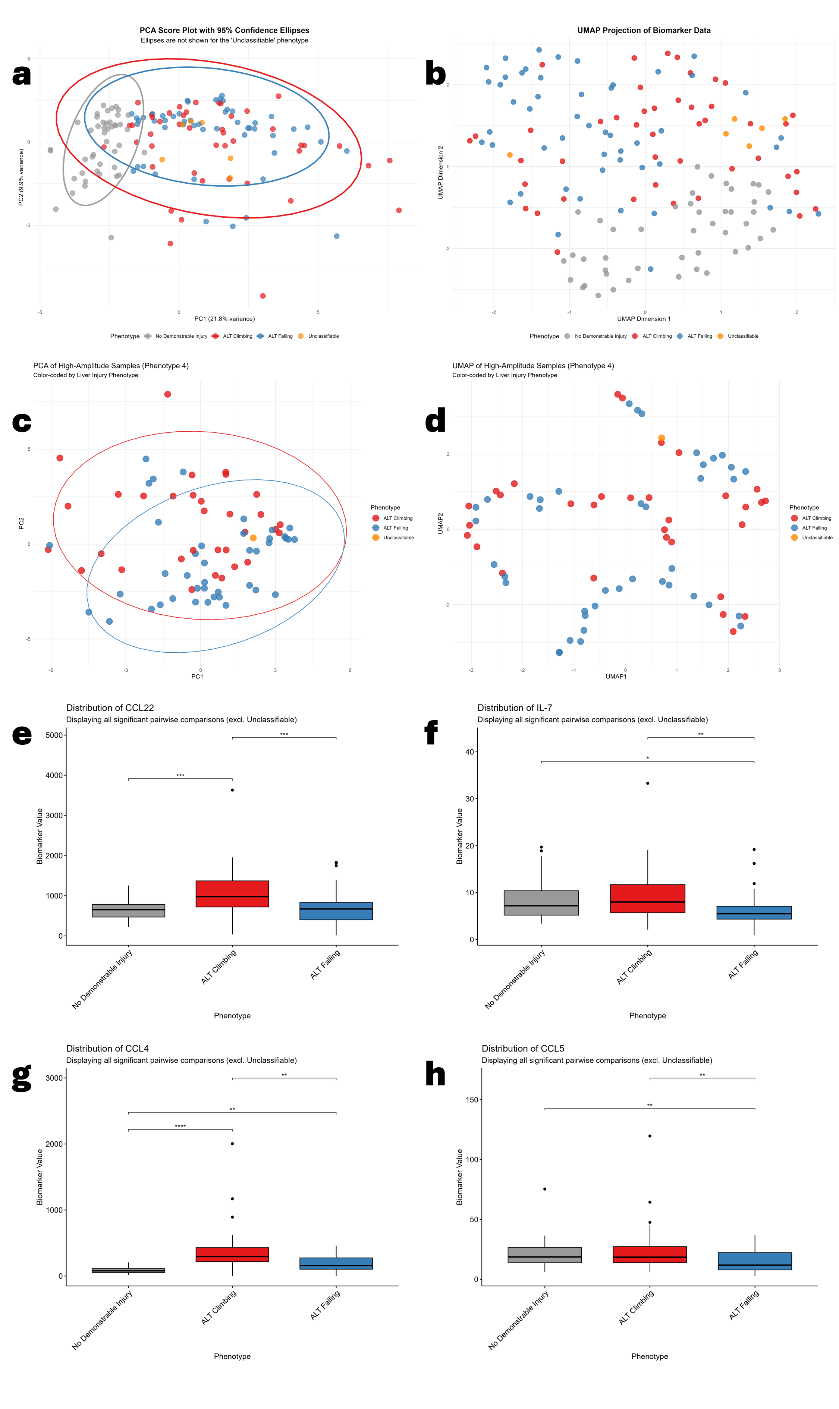

**Fig. S3-3. A) Principal Component Analysis PC1 vs. PC2, showing that dimensionality reduction can distinguish injury from non-injury, but fails to distinguish subsequent ALT trajectory. B) UMAP analysis, demonstrating the same clustering behaviour as in A. C-D) Principal Component and UMAP analyses, restricted to samples following the development of ALT>1000 demonstrated the same failure to distinguish subsequent ALT trajectory as in A and B. E-H) Kruskal-Wallis testing demonstrated that CCL22, IL-7, CCL4, CCL5 distinguished ALT-climbing vs. ALT-falling samples.**

| **Biomarker** | **p-value** |
| --- | --- |
| IL-7 | 0.000615 |
| WCC | 0.00089 |
| CCL22 | 0.000978 |
| CCL4 | 0.001039 |
| IL-12p70 | 0.001739 |
| HMGB1 | 0.005824 |
| Bilirubin | 0.006093 |
| IL-15 | 0.007002 |
| Osteopontin | 0.009147 |
| IL-9 | 0.011045 |
| CCL13 | 0.015601 |
| CCL5 | 0.016281 |
| CCL3 | 0.018422 |
| IL-10 | 0.01897 |
| miR-122 | 0.019275 |
| CCL26 | 0.037036 |

**Table S3-4. Biomarkers found to be significantly different between ALT climbing and ALT falling phenotypes in hepatotoxicity APAP DILI samples ordered by p-value.**

#### S4. Development and validation of predictive models for ALT trajectory

###### S4.1 Detailed modelling methods

To find the best combination of biomarkers for our predictive model, we sought to balance predictive power with the risk of overfitting. Our goal was to find the smallest set of biomarkers that provided the best performance.

For models with one, two, three, and four biomarkers, we tested every possible combination in an exhaustive search of 378,658 potential models. As this approach was computationally impractical for larger models, we used a more targeted beam search strategy for models of five to eight biomarkers. This approach iteratively builds larger models by taking the 500 best-performing models at the previous combination size (e.g. n=4) and testing the addition of every other available biomarker. After establishing the optimal correlation threshold of ρ<0.5 (see S6), we re-derived our models using the same strategy but expanded the beam search to the top 1000 models at each step (321,682 total models tested).

###### S4.2 Exploratory analysis of individual and paired biomarkers

Before building larger models, we first evaluated the ability of individual biomarkers and simple pairwise combinations to predict ALT trajectory. Using a Kernel Naive Bayes model, we found that eighteen individual biomarkers outperformed ALT alone. The performance of the top nine individual biomarkers is shown in Table S4-1.

We next tested pairwise combinations, excluding pairs with a Spearman’s correlation of ρ≥0.7 to prioritize biologically distinct signals. Combinations of biomarkers consistently improved classification ability, illustrating the value of a multi-dimensional approach (Table S4-1).

| Biomarker | Derivation AUROC | Best combination | Absolute Spearman correlation | Combination derivation AUROC |
| --- | --- | --- | --- | --- |
| IL-10 | 0.783 | CCL4 | 0.078 | 0.900 |
| CCL5 | 0.783 | IL-9 | 0.180 | 0.861 |
| WBC | 0.781 | IL-7 | 0.311 | 0.836 |
| HMGB1 | 0.774 | SLPI | 0.199 | 0.878 |
| CCL2 (Luminex) | 0.767 | IL-9 | 0.050 | 0.839 |
| IL-7 | 0.761 | WBC | 0.311 | 0.836 |
| Osteopontin | 0.714 | IL-7 | 0.268 | 0.836 |
| CCL3 | 0.708 | IL-10 | 0.360 | 0.836 |
| Bilirubin | 0.700 | CCL5 | 0.347 | 0.844 |
| IL-17A | 0.699 | IL-10 | 0.099 | 0.867 |
| CCL22 | 0.683 | WBC | 0.040 | 0.783 |
| IL-9 | 0.681 | CCL5 | 0.180 | 0.861 |
| C3a:C3 ratio | 0.675 | WBC | 0.131 | 0.792 |
| IL-2 | 0.675 | IL-10 | 0.046 | 0.842 |
| K18 | 0.669 | IL-10 | 0.648 | 0.872 |
| CCL4 | 0.658 | IL-10 | 0.078 | 0.900 |
| SLPI | 0.656 | HMGB1 | 0.199 | 0.878 |
| CSF3 | 0.653 | IL-10 | 0.389 | 0.839 |

**Table S4-1. Eighteen individual biomarkers outperformed ALT (AUC = 0.625) for the prediction of the future trajectory of ALT. The impact of combining an additional biomarker is shown (best performing combination for each).**

**Fig. S4-2: Incremental gains to AUROC when additional biomarkers are integrated.**

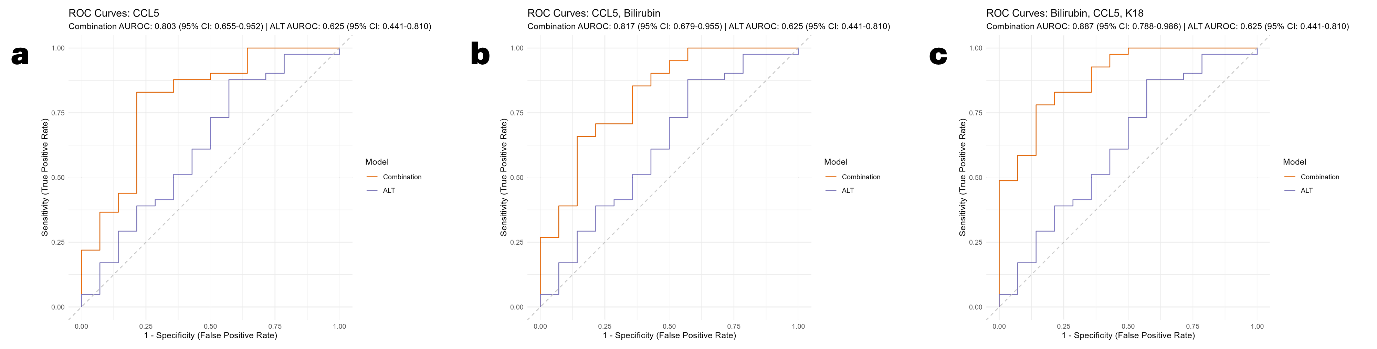

**Derived ROC curve for the prediction of ALT trajectory for ALT only (blue) and A) CCL5, B) CCL5+bilirubin, C) CCL5, bilirubin and K18; demonstrating the incremental increase with each additional biomarker to the model.**

###### S4.3 Model Validation Details

Validation was performed on an independent cohort where cytokine analysis was conducted exclusively using the Luminex Discovery platform. To avoid confounding from inter-platform variability in quantification, we restricted the validation analysis to models constructed solely from the 15 biomarkers quantified using identical platforms in both the derivation and validation cohorts. These biomarkers were: HMGB1, K18, miR-122, CCL2, CCL5, MCSFR, alkaline phosphatase, bilirubin, haemoglobin, creatinine, white cell count (WCC), urea, potassium, sodium, and INR. This generated 12,910 possible combinations (of 1:8 biomarkers) of which 1,929 met the correlation threshold. Comparison of combination sizes larger than 8 was not feasible (9-biomarker combinations only generated 6 options). The rapid decline suggests the number of potential combinations fell primarily due to the correlation threshold, but symmetry will also play a role in decreasing numbers as model sizes increase (i.e. choosing a small number of items to include is the same as choosing a small number of items to exclude, and therefore the number of possible combinations peaks in the middle and then symmetrically decreases).

###### S4.4. Model performance with absolute biomarker correlations <0.7

**Fig. S4-3: Model performance with less restricted correlation.**

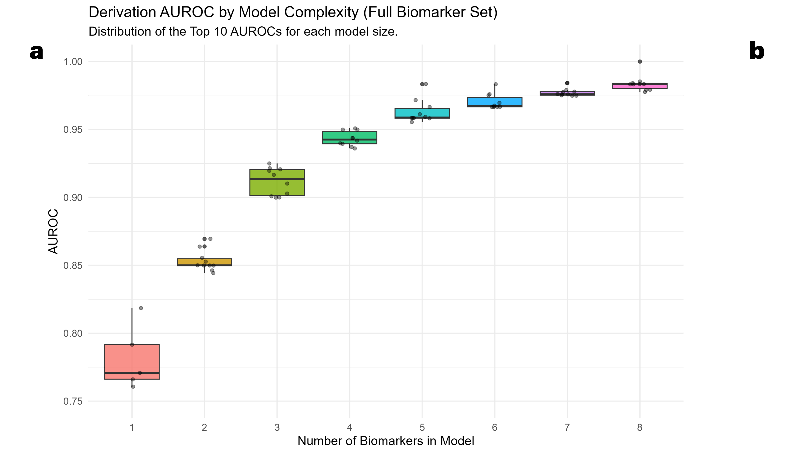

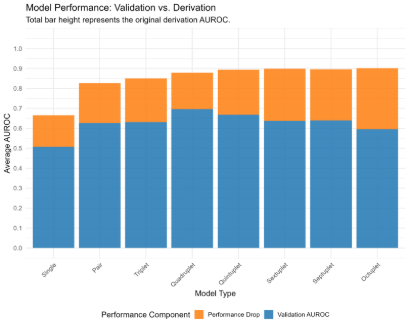

**A) Boxplot showing AUC of the top 10 models when absolute correlations are limited to <0.7. An inflection point is seen at 4 biomarkers, suggesting a risk of overfitting beyond this point. B) Stacked bar chart comparing the performance of the top 10 restricted biomarker set models, demonstrating that models with >4 biomarkers suffered from over-fitting at validation.**

#### S5. Optimising model performance by ensuring biomarkers are mechanistically distinct

Sensitivity analysis for absolute biomarker correlation thresholds was undertaken by examining the performance of models in the validation cohort across each number of biomarkers and a range of correlation thresholds, to assess both the impact on the AUC of the top 10 models for each number of biomarkers (Fig. S5-1A) and the impact on the fall in AUC passing from validation to derivation stages (Fig. S5-1B). We identified that a threshold of 0.5 appeared to achieve the best compromise between validation model performance and the translation of models from derivation to validation.

**Fig. S6-1: Sensitivity analysis of biomarker combination size and permitted maximum biomarker correlation.**
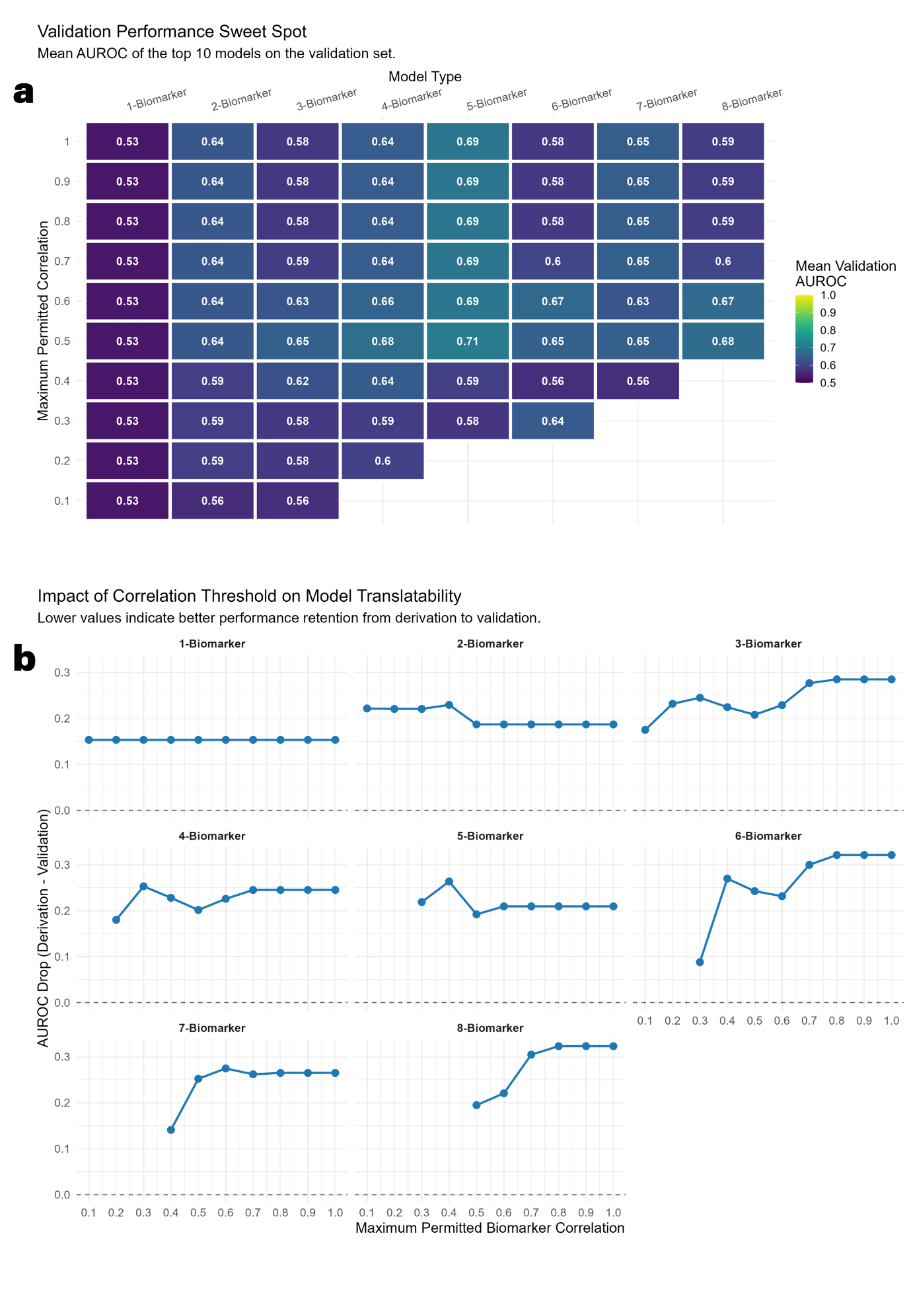

**A) Heatmap showing the mean performance of the top 10 models at validation for every feasible combination of maximum permitted biomarker correlation and model complexity. B) Facet plot showing the performance drop seen from derivation to validation at each level of maximum permitted biomarker correlation.**

#### S6. Biomarkers analysed for study

| **Biomarker** | **Method** | **Obtained during routine clinical care?** | **Used in exploratory derivation cohort?** | **Used in final derivation and validation cohort?** |
| --- | --- | --- | --- | --- |
| Alkaline Phosphatase (ALP) | Clinical labs | Yes | Yes | Yes |
| ALT | Clinical labs | Yes | As reference | As reference |
| Antileukoproteinase(SLPI) | Mesoscale Discovery R-PLEX (HUMAN) | No | Yes | No |
| Bilirubin | Clinical labs | Yes | Yes | Yes |
| C3 | Mesoscale Discovery R-PLEX (HUMAN) | No | Yes | No |
| C3a | Mesoscale Discovery R-PLEX (HUMAN) | No | Yes | No |
| C3a:C3 ratio | Derived from other values | No | Yes | No |
| CCL1 | Mesoscale Discovery U-PLEX (HUMAN) | No | Yes | No |
| CCL2 | Mesoscale Discovery V-PLEX (HUMAN) | No | Yes | No |
| CCL2 | Luminex Human Discovery Assay | No | Yes | Yes |
| CCL3 | Mesoscale Discovery V-PLEX (HUMAN) | No | Yes | No |
| CCL4 | Mesoscale Discovery V-PLEX (HUMAN) | No | Yes | No |
| CCL5 | Luminex Human Discovery Assay | No | Yes | Yes |
| CCL8 | Luminex Human Discovery Assay | No | No | No |
| CCL11 | Mesoscale Discovery V-PLEX (HUMAN) | No | Yes | No |
| CCL13 | Mesoscale Discovery V-PLEX (HUMAN) | No | Yes | No |
| CCL26 | Mesoscale Discovery V-PLEX (HUMAN) | No | Yes | No |
| Creatinine | Clinical labs | Yes | Yes | Yes |
| CRP | Mesoscale Discovery V-PLEX (HUMAN) | No | Yes | No |
| CSF1 | Mesoscale Discovery U-PLEX (HUMAN) | No | Yes | No |
| CSF2 | Mesoscale Discovery V-PLEX (HUMAN) | No | Yes | No |
| CSF3 | Mesoscale Discovery U-PLEX (HUMAN) | No | Yes | No |
| CX3CL1 | Luminex Human Discovery Assay | No | No | No |
| CXCL1 | Mesoscale Discovery U-PLEX (HUMAN) | No | Yes | No |
| CXCL9 | Mesoscale Discovery U-PLEX (HUMAN) | No | Yes | No |
| CXCL10 | Mesoscale Discovery V-PLEX (HUMAN) | No | Yes | No |
| CXCL14 | Luminex Human Discovery Assay | No | No | No |
| Haemoglobin | Clinical labs | Yes | Yes | Yes |
| HMGB1 | IBL-Tecan Express ELISA | No | Yes | Yes |
| IFN-γ | Mesoscale Discovery V-PLEX (HUMAN) | No | Yes | No |
| IL-1a | Mesoscale Discovery V-PLEX (HUMAN) | No | No | No |
| IL-1β | Mesoscale Discovery V-PLEX (HUMAN) | No | No | No |
| IL-2 | Mesoscale Discovery V-PLEX (HUMAN) | No | Yes | No |
| IL-3 | Mesoscale Discovery V-PLEX (HUMAN) | No | No | No |
| IL-4 | Mesoscale Discovery V-PLEX (HUMAN) | No | No | No |
| IL-6 | Mesoscale Discovery V-PLEX (HUMAN) | No | Yes | No |
| IL-7 | Mesoscale Discovery V-PLEX (HUMAN) | No | Yes | No |
| IL-8 | Mesoscale Discovery V-PLEX (HUMAN) | No | Yes | No |
| IL-9 | Mesoscale Discovery V-PLEX (HUMAN) | No | Yes | No |
| IL-10 | Mesoscale Discovery V-PLEX (HUMAN) | No | Yes | No |
| IL-12p70 | Mesoscale Discovery V-PLEX (HUMAN) | No | Yes | No |
| IL-13 | Mesoscale Discovery V-PLEX (HUMAN) | No | Yes | No |
| IL-15 | Mesoscale Discovery V-PLEX (HUMAN) | No | Yes | No |
| IL-17A | Mesoscale Discovery V-PLEX (HUMAN) | No | Yes | No |
| IL-18 | Mesoscale Discovery U-PLEX (HUMAN) | No | Yes | No |
| IL-21 | Mesoscale Discovery V-PLEX (HUMAN) | No | No | No |
| IL-22 | Mesoscale Discovery V-PLEX (HUMAN) | No | Yes | No |
| IL-23 | Mesoscale Discovery V-PLEX (HUMAN) | No | No | No |
| INR | Clinical labs | Yes | Yes | Yes |
| K18 | Peviva M65 ELISA | No | Yes | Yes |
| MCSFR | Luminex Human Discovery Assay | No | Yes | Yes |
| miR-122 | RT qPCR. Normalised to miR-39 pre-extraction spike-in. | No | Yes | Yes |
| Osteopontin | Luminex Human Discovery Assay | No | Yes | No |
| PDGF-BB | Luminex Human Discovery Assay | No | Yes | No |
| Potassium | Clinical labs | Yes | Yes | Yes |
| sICAM-1 (CD54) | Mesoscale Discovery V-PLEX (HUMAN) | No | Yes | No |
| Sodium | Clinical labs | Yes | Yes | Yes |
| sVCAM-1 (CD106) | Mesoscale Discovery V-PLEX (HUMAN) | No | Yes | No |
| TNF-α | Mesoscale Discovery V-PLEX (HUMAN) | No | Yes | No |
| TNF-β | Mesoscale Discovery V-PLEX (HUMAN) | No | Yes | No |
| Urea | Clinical labs | Yes | Yes | Yes |
| VEGF-A | Mesoscale Discovery V-PLEX (HUMAN) | No | Yes | No |
| White Blood Cell count (WBC, WCC) | Clinical labs | Yes | Yes | Yes |
